# Clinical and epidemiological characteristics of Coronavirus Disease 2019 (COVID-19) patients

**DOI:** 10.1101/2020.04.02.20050989

**Authors:** Shing Cheng Tan

## Abstract

**Background:** Numerous groups have reported the clinical and epidemiological characteristics of Coronavirus Disease 2019 (COVID-19) cases; however, the data remained inconsistent. This paper aimed to pool the available data to provide a more complete picture of the characteristics of COVID-19 patients.

**Methods:** A systematic review and pooled analysis was performed. Eligible studies were identified from database and hand searches up to March 2, 2020. Data on clinical (including laboratory and radiological) and epidemiological (including demographic) characteristics of confirmed COVID-19 cases were extracted and combined by simple pooling.

**Results:** Of 644 studies identified, 69 studies (involving 48,926 patients) were included in the analysis. The average age of the patients was 49.16 years. A total of 51.46% of the patients were men and 52.32% were non-smokers. Hypertension (50.82%) and diabetes (20.89%) were the most frequent comorbidities observed. The most common symptoms were fever (83.21%), cough (61.74%), and myalgia or fatigue (30.22%). Altered levels of blood and biochemical parameters were observed in a proportion of the patients. Most of the patients (78.50%) had bilateral lung involvements, and 5.86% showed no CT findings indicative of viral pneumonia. Acute respiratory distress syndrome (28.36%), acute cardiac injury (7.89%) and acute kidney injury (7.60%) were the most common complications recorded.

**Conclusions:** Clinical and epidemiological characteristics of COVID-19 patients were mostly heterogeneous and non-specific. This is the most comprehensive report of the characteristics of COVID-19 patients to date. The information presented is important for improving our understanding of the spectrum and impact of this novel disease.

## Introduction

There is an ongoing pandemic of viral pneumonia called Coronavirus Disease 2019 (COVID-19) which is caused by severe acute respiratory syndrome coronavirus 2 (SARS-CoV-2) infection. At the time of this writing, the disease has been reported to affect 634,835 people in more than 200 countries, territories or areas, and cause 29,891 deaths (1). Understanding the clinical and epidemiological characteristics of the disease is important for informing public health decision making, which would enable improvement of surveillance and effective planning of treatment. In January 2020, Huang et al. (2) published the first report on the characteristics of a series of COVID-19 patients in Wuhan, China, the first epicenter of the outbreak. Following that, many other groups have reported the clinical and epidemiological features of COVID-19 patients, both in China and in other parts of the world (3-12). However, most of these reports were limited by a small sample size, and the characteristics reported appeared to be inconsistent. For example, in a report involving 99 patients, Chen et al. (3) noted a much higher proportion of men than women, and suggested that men were generally more susceptible to SARS-CoV-2 infection. On the other hand, Shi et al. (4) found that the male-to-female ratio among the 81 patients included was close to 1:1, indicating that both genders were equally susceptible to COVID-19. Recently, the World Health Organization (WHO) has declared COVID-19 a global pandemic and the contagion shows no sign of slowing down (13). Thus, it is a timely prompt to obtain a more precise understanding of the disease by combining all available data in the literature. In this study, a systematic review and pooled analysis was performed to characterize the clinical and epidemiological features of COVID-19 patients.

## Methods

### Search strategy and selection criteria

Three separate searches were performed on PubMed database on March 2, 2020, using the keywords “COVID-19”, “2019-nCoV” and “SARS-CoV-2”. No language restriction was applied. After removal of duplicated records, screening by title and abstract was performed to identify potentially relevant studies. The full-texts of these potentially relevant studies were then evaluated. The reference lists of these studies were also hand-searched to identify additional records. Studies were included if they reported any clinical and/or epidemiological data of confirmed COVID-19 patients, regardless of their study design. However, review papers or studies employing secondary analysis of the previously available data were excluded from the analysis. In case of overlapping studies, the ones which the largest sample size or the most complete data set were included.

### Data analysis

The following data were extracted from the included studies: name of first author, country, date of diagnosis, demographic data, smoking status, comorbidities, signs and symptoms, laboratory/biochemical data, CT findings, and complications of the disease. A simple pooling of data was performed to provide an overall summary of the clinical and epidemiological characteristics of the patients. All data are reported as absolute number and/or mean. As patient-level data were not available in majority of the studies, standard deviation could not be calculated.

### Role of the funding source

There was no funding source for this study. The corresponding author had full access to all the data in the study and had final responsibility for the decision to submit for publication.

## Results

The flowchart of study selection is depicted in Figure 1. A total of 644 records were identified from the PubMed searches. After deduplication, 517 unique records were identified. Following screening by title and abstract, 440 records were removed and full-texts of the remaining 77 studies were retrieved for assessment of eligibility. Five records were subsequently removed as they reported secondary data or contained insufficient data on clinical and/or epidemiological characteristics of the patients. In addition, three overlapping records were excluded. Eventually, 69 studies were included in the current analysis (2-12, 14-71). These 69 studies comprised a total of 48,926 confirmed COVID-19 patients, although only appropriate subcohorts were used in the analysis of each clinical/epidemiological characteristic. A great majority of these studies were from mainland China, while five were from Korea and one each from the USA, Germany, France, Australia, Italy, Singapore, Vietnam, Nepal, Hong Kong, and Taiwan.

**Figure 1:**
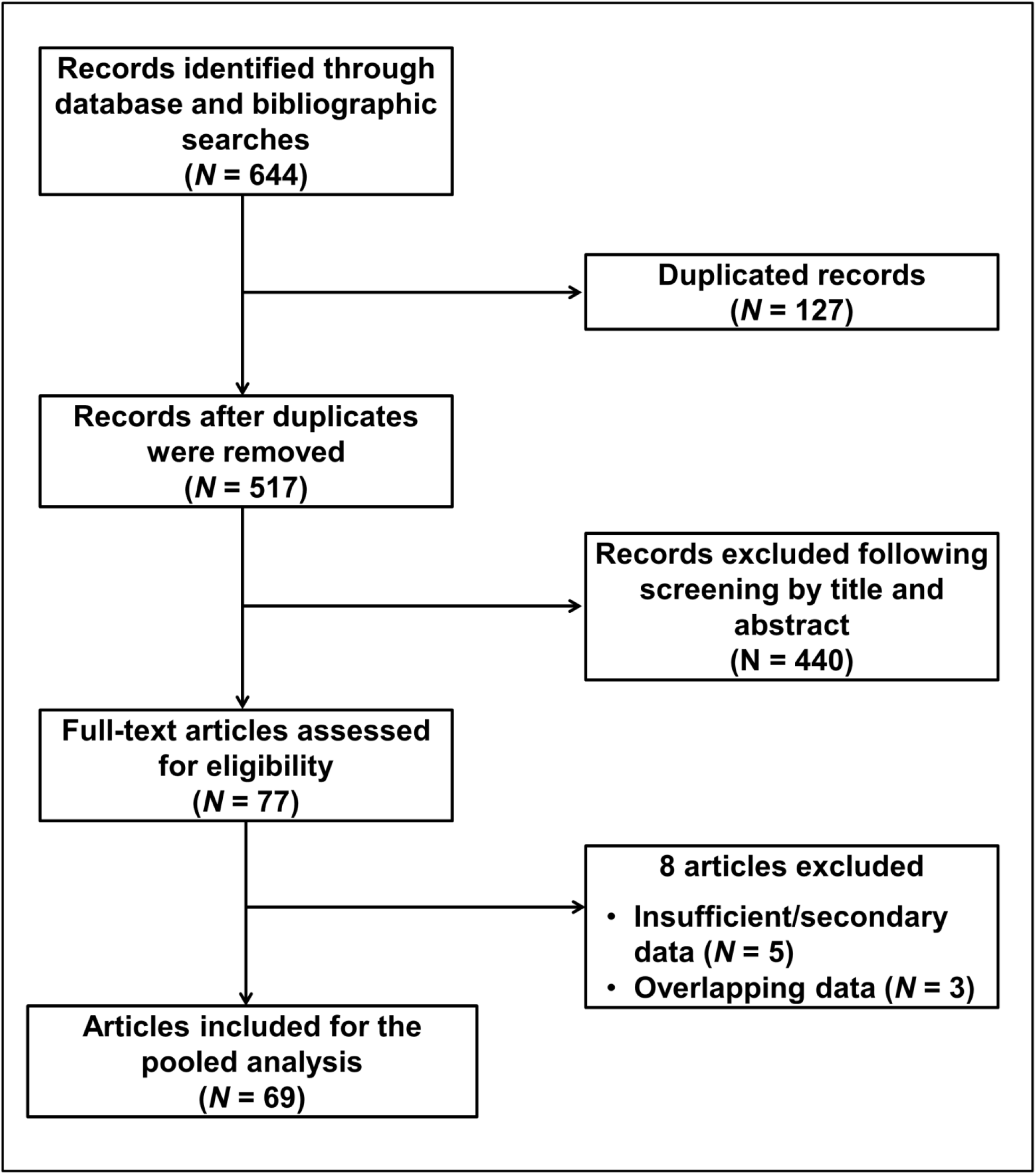
Flow chart of study selection

The characteristics of the patients are shown in Table 1. The mean age of the patients was 49.16 years. The number of male patients was slightly higher than that of female (51.46% vs. 48.54%), and the number of non-smokers was slightly higher than that of smokers (52.32% vs. 47.68%). A minority (25.93%) of the patients had comorbidities, among which hypertension was the most common (50.82%), followed by diabetes (20.89%), cardiovascular and cerebrovascular diseases (16.54%), respiratory system disease (9.70%) and malignancy (2.05%). Most of the patients showed at least one sign or symptom at presentation, with only 0.8% of the patients were asymptomatic. The most common symptoms seen included fever (83.21%), cough (61.74%), myalgia or fatigue (30.22%), sputum production (20.22%), and dyspnea (16.97%). On average, the length from illness onset to dyspnea was 4.99 days.

**Table 1:** Characteristics of COVID-19 patients

| Characteristics | Number of patients analyzed | Number/Mean |
| --- | --- | --- |
| Age (years) | 1,725 | 49.16 |
| Sex | 44,760 |  |
| Male |  | 23,032 (51.46%) |
| Female |  | 21,728 (48.54%) |
| Smoking status | 2,030 |  |
| Smokers |  | 968 (47.68%) |
| Non-smokers |  | 1,062 (52.32%) |
| Comorbidities | 21,020 |  |
| Yes |  | 5,451 (25.93%) |
| No |  | 15,569 (74.07%) |
| Type of comorbidities | 5,450 |  |
| Hypertension |  | 2,683 (50.82%) |
| Diabetes |  | 1,103 (20.89%) |
| Cardiovascular and cerebrovascular diseases |  | 873 (16.54%) |
| Respiratory system disease |  | 512 (9.70%) |
| Malignancy |  | 108 (2.05%) |
| Presence of symptoms | 3,117 |  |
| Yes |  | 3092 (99.20%) |
| No |  | 25 (0.80%) |
| Signs and symptoms |  |  |
| Fever | 3,097 | 2577 (83.21%) |
| Cough | 3,097 | 1912 (61.74%) |
| Myalgia or fatigue | 3,117 | 942 (30.22%) |
| Sputum production | 3,115 | 630 (20.22%) |
| Dyspnea | 3,093 | 525 (16.97%) |
| Length from illness onset to dyspnea (days) | 179 | 4.99 |
| Headache | 3,114 | 325 (10.44%) |
| Sore throat | 3,117 | 250 (8.02%) |
| Chill | 3,117 | 201 (6.45%) |
| Nausea and vomiting | 3,116 | 142 (4.56%) |
| Diarrhea | 3,113 | 136 (4.37%) |
| Chest tightness but not pain | 3,117 | 94 (3.02%) |
| Nasal congestion or runny nose | 3,117 | 64 (2.05%) |
| Loss of appetite | 3,116 | 62 (1.99%) |
| Abdominal pain | 3,116 | 38 (1.22%) |
| Rhinorrhoea | 3,117 | 35 (1.12%) |
| Pharyngalgia | 3,117 | 33 (1.06%) |
| Dizziness | 3,117 | 32 (1.03%) |
| Chest pain | 3,117 | 25 (0.80%) |
| Hemoptysis | 3,115 | 23 (0.74%) |
| Respiratory rate >24 breaths per min | 3,117 | 22 (0.71%) |
| Malaise | 3,117 | 21 (0.67%) |
| Heart palpitation | 3,117 | 14 (0.45%) |
| Conjunctivitis | 3,117 | 10 (0.32%) |
| Confusion | 3,117 | 10 (0.32%) |
| Blood in sputum | 3,117 | 3 (0.10%) |
| Anthralgia | 3,117 | 2 (0.06%) |
| Gastrointestinal reaction | 3,117 | 1 (0.03%) |
| Back pain | 3,117 | 1 (0.03%) |
| Constipation | 3,117 | 1 (0.03%) |
| Skin tingling | 3,117 | 1 (0.03%) |
| Somnolence | 3,117 | 1 (0.03%) |
| Sneezing | 3,117 | 1 (0.03%) |
| Jaundice | 3,117 | 1 (0.03%) |
| Respiratory symptoms* | 3,117 | 7 (0.22%) |
| Laboratory findings |  |  |
| Leukocyte count ($\times 10^9$ per L) | 801 | 5.60 |
| Decreased | 961 | 232 (24.14%) |
| Normal | 961 | 606 (63.06%) |
| Increased | 961 | 123 (12.80%) |
| Neutrophil count ($\times 10^9$ per L) | 571 | 3.63 |
| Lymphocyte count ($\times 10^9$ per L) | 849 | 0.97 |
| Decreased | 827 | 425 (51.39%) |
| Normal | 827 | 402 (48.61%) |
| Hemoglobin, g/L | 336 | 127.47 |
| Platelet count, ($10^9$ per L) | 635 | 181.58 |
| Decreased | 561 | 54 (9.63%) |
| Prothrombin time (s) | 562 | 11.94 |
| Activated partial thromboplastin time (s) | 360 | 29.96 |
| D-dimer (mg/L) | 726 | 1.13 |
| D-dimer (mg/L) when one study was removed<sup>†</sup> | 645 | 0.45 |
| Albumin (g/L) | 473 | 36.84 |
| Alanine aminotransferase (U/L) | 634 | 31.55 |
| Aspartate aminotransferase (U/L) | 614 | 32.59 |
| Normal | 499 | 362 (72.55%) |
| Increased | 499 | 137 (27.45%) |
| Total bilirubin (mmol/L) | 659 | 12.04 |
| Potassium (mmol/L) | 44 | 4.23 |
| Sodium (mmol/L) | 44 | 138.48 |
| Creatinine (μmol/L) | 633 | 72.45 |
| Normal | 231 | 219 (94.81%) |
| Increased | 231 | 12 (5.19%) |
| Creatine kinase (U/L) | 636 | 93.66 |
| Normal | 400 | 347 (86.75%) |
| Increased | 400 | 53 (13.25%) |
| Lactate dehydrogenase (U/L) | 547 | 261.44 |
| Normal | 413 | 224 (54.24%) |
| Increased | 413 | 189 (45.76%) |
| Hypersensitive troponin I (pg/mL) | 179 | 5.91 |
| >28 (99th percentile) | 28 | 1 (3.57%) |
| Procalcitonin (ng/mL) | 334 | 0.21 |
| Normal | 359 | 111 (30.92%) |
| Increased | 359 | 248 (69.08%) |
| Increased C-reactive protein | 870 | 629 (72.30%) |
| CT findings |  |  |
| Lung involvement | 1,144 |  |
| Bilateral |  | 898 (78.50%) |
| Unilateral |  | 179 (15.65%) |
| No abnormalities |  | 67 (5.86%) |
| Number of lobes affected | 323 |  |
| 1 |  | 55 (17.03%) |
| 2 |  | 37 (11.46%) |
| 3 |  | 41 (12.69%) |
| 4 |  | 57 (17.65%) |
| 5 |  | 133 (41.18%) |
| Complications | 342 |  |
| Acute respiratory distress syndrome |  | 97 (28.36%) |
| Acute cardiac injury |  | 27 (7.89%) |
| Acute kidney injury |  | 26 (7.60%) |
| Arrhythmia |  | 23 (6.73%) |
| Shock |  | 20 (5.85%) |
| Hyperglycemia |  | 18 (5.26%) |
| Hepatic insufficiency / liver dysfunction |  | 17 (4.97%) |
| Acute respiratory injury |  | 8 (2.34%) |
| Hospital acquired/Ventilator associated pneumonia |  | 8 (2.34%) |
| RNAemia |  | 6 (1.75%) |
| Secondary infection |  | 4 (1.17%) |
| Renal insufficiency |  | 2 (0.58%) |
| Gastrointestinal hemorrhage |  | 2 (0.58%) |
| Cardiac failure |  | 1 (0.29%) |
| Pneumothorax |  | 1 (0.29%) |
| Urinary tract infection |  | 1 (0.29%) |
| Bacteremia |  | 1 (0.29%) |

\* Respiratory symptoms included multiple symptoms such as cough and sore throat, but they were not reported separately in the original publications
† Data from Shi et al. (4) was removed as the value reported was an outlier

The laboratory findings of the patients are also shown in Table 1. Majority (63.06%) of the patients had a normal leukocyte count, but as much as 24.14% and 12.80% respectively showed decreased and increased leukocyte counts. Besides, 51.39% of the patients had a decreased lymphocyte count and 9.63% had a decreased platelet count. Increased levels of aspartate aminotransferase, creatinine, creatine kinase, lactate dehydrogenase, hypersensitive troponin I, procalcitonin and C-reactive protein were observed in 27.45%, 5.19%, 13.25%, 45.76%, 3.57%, 69.08% and 72.30% of the patients, respectively.

Analysis of computed tomography (CT) data revealed that most of the patients (78.50%) had bilateral lung involvements, while 15.65% had unilateral lung involvements and 5.86% showed no sign of viral pneumonia. In 41.18% of the patients, all five lung lobes were affected. On the other hand, as much as 17.03%, 11.46%, 12.69% and 17.65% patients respectively had one, two, three and four lobes affected.

It was unknown how many patients had complications following SARS-CoV-2 infection. Nonetheless, among the wide spectrum of complications observed, acute respiratory distress syndrome (ARDS) was most frequently documented (28.36%), followed by acute cardiac injury (7.89%) and acute kidney injury (7.60%).

## Discussion

COVID-19 poses a significant burden on the healthcare system all over the world. A complete understanding of the characteristics of the disease is important for effective surveillance and public health response measures to be implemented in a timely manner. Currently, although we have some basic understanding of the clinical and epidemiological features of COVID-19 patients, our knowledge is insufficient. This is because inconsistencies still exist in the findings of many published reports, and the sample sizes in most of these reports were too small for a reliable summary to be made. In this work, a systematic review and pooled analysis was performed to combine data from 69 previous reports, in order to yield a more accurate summary of the clinical and epidemiological characteristics of COVID-19 patients.

In many instances, susceptibility to viral infections may be related to factors such as gender and smoking habits (72-74). For the former, it is believed that X chromosome inactivation in females may cause cellular mosaicism which ensures the presence of at least one functional copy of X-linked immune genes, thus conferring women an increased resistance against viral infections (75). In addition, estrogen, the major female sex hormone, is known to promote adaptive immune response (76), while testosterone, the primary sex hormone in men, could contribute to the suppression of the innate immune response, rendering men more susceptible to viral infections (77, 78). On the other hand, cigarette smoking may reduce the level of circulating immunoglobulins, immune cells, and pro-inflammatory cytokines, as well as disrupt the response of antibodies to antigens (72). For these reasons, some studies have suggested that men and smokers are more susceptible to SARS-CoV-2 infections (3, 79). In the present work, we noted that the ratios of male to female and smokers to non-smokers were close to 1:1. Although the relative risk or odds ratio of the association between these variables and SARS-CoV-2 infection could not be computed due to the lack of a comparison group, a proportion of approximately 1:1 suggests that susceptibility to SARS-CoV-2 infection is universal.

The present work also showed that the symptoms of COVID-19 were generally non-specific, thus the disease cannot be reliably distinguished from other infectious diseases based on the symptoms alone. The most commonly observed symptoms were similar to those of the previous coronavirus disease outbreaks (MERS and SARS), i.e. fever, cough, and myalgia or fatigue. Nevertheless, compared to MERS and SARS, the proportion of afebrile patients was much higher in COVID-19 (16.79% cf. 2% in MERS and 0-1% in SARS) (80). This indicates that a significant number of COVID-19 patients would be missed if surveillance and monitoring systems focus largely on temperature screening, as commonly practiced in airports (81, 82). The present work also showed that dyspnea was observed in only 16.97% of the patients. This contradicts with advisories and guidelines published by many health authorities, which suggest that dyspnea is a commonly observed symptom in COVID-19 (83-89). Besides, it was observed that 0.80% of the patients were asymptomatic. Currently, whether asymptomatic patients can transmit the virus to other individuals is not fully known, but it is highly possible (35). Thus, although the number of asymptomatic patients was low, identifying and isolating such patients to prevent uncontrolled disease spread would prove very challenging. It is therefore important for diagnostic tests to be performed on asymptomatic medium- and high-risk individuals to facilitate early detection and prevention of SARS-CoV-2 transmission. In addition to nucleic acid testing using real-time reverse transcription polymerase chain reaction, some studies have suggested the potential usability of chest CT for COVID-19 diagnosis (90, 91). However, in the present work, we found that 5.86% of the patients did not have abnormalities on CT scans. Thus, although CT findings have substantial accuracy in identifying the disease (90, 91), the results need to be interpreted with caution.

SARS-CoV-2 is known to infect a cell by first binding to its angiotensin converting enzyme 2 (ACE2) receptor (92). Apart from the lung, high expression of ACE2 receptor is also observed in several other organs such as the heart, kidney, and intestine, as well as the in lymphocytes (93, 94). Several previous studies reported that a decreased lymphocyte count was a common feature of SARS-CoV-2 infection (2, 3, 7, 30, 41). In the present work, lymphopenia was observed in 51.39% of the patients. A decreased lymphocyte count implies a weakening adaptive immune system. Considering the high expression of ACE2 in lymphocytes, it has been postulated that SARS-CoV-2 may directly infect and attack lymphocytes, thus impairing the immune system (95). Besides the decrease in lymphocyte count, the present study also showed that many COVID-19 patients had increased levels of C-reactive protein, creatine kinase, lactate dehydrogenase and procalcitonin. High levels of C-reactive protein and creatine kinase suggest that sustained inflammatory response occurs following SARS-CoV-2 infection, whereas high levels of lactate dehydrogenase and procalcitonin indicate the virus could, either directly or indirectly, cause tissue injury. A previous work reported that a low lymphocyte count and high levels of lactate dehydrogenase and C-reactive protein were among the parameters found to be correlated with severity of lung injury in COVID-19 patients, as measured using the Murray scores (17). Nonetheless, the relationship between these variables and severity of COVID-19 was not investigated in the present work due to the insufficient data available.

A few limitations exist in the present work. First, there were instances where a same patient was described in multiple reports (15, 96). Efforts have been made to identify such patients by meticulously reviewing the descriptions of the patients in each report. However, since all patients were anonymized, it was challenging to identify all overlapping patients. Thus, there is a possibility that some overlapping patients were not removed from our analysis and their characteristics were overreported. Besides, since patient-level data were not reported in most of the studies, median values and standard deviations, which understandably provide more meaningful information, could not be computed. Finally, as mentioned above, various comparisons among the patients (e.g. severe vs. mild, and death vs. survivor) could not be analyzed due to insufficient data available.

## Conclusion

In conclusion, this report has successfully provided a more complete picture of the clinical and epidemiological characteristics of COVID-19 patients. A wide variation exists in the clinical manifestation of the disease. As the outbreak continues to escalate and SARS-CoV-2 continues to mutate (97), it is important to consistently update the characteristics of the patients in order to monitor whether evolving strains of the virus could cause the disease differently. Sharing of clinical and epidemiological data among the scientific community is highly important for informing public health decision making for controlling the spread of the disease.

## Data Availability

The data that support the findings of this study are available from the corresponding author upon reasonable request.

## Contributor

SCT contributed solely to this work.

## Declaration of interests

The author declares no competing interests.

## Acknowledgments

No funding was obtained for this work.

## Notes

### Competing Interest Statement

The authors have declared no competing interest.

